# Blood Pressure and Risk of Clinical Events in Peripheral Artery Disease

**DOI:** 10.1101/2025.09.29.25336935

**Authors:** Saket Girotra, Mary Vaughan-Sarrazin, James A. de Lemos, Paul S. Chan, Ajay K. Gupta, Qiang Li, Cathy Nguyen, Brian C. Lund, Rohit Nathani, Richard M. Hoffman, Glenn T. Gobbel, Mohammed Al-Garadi, Michael E. Matheny, Dharam J. Kumbhani, Anthony A. Bavry, Shipra Arya, Kim G. Smolderen, Diana Jalal, Wanpen Vongpatanasin, Joshua A. Beckman

## Abstract

**Background:** Current guidelines recommend a blood pressure (BP) goal ≤130/80 mm Hg in peripheral artery disease (PAD) even though few PAD patients were enrolled in the landmark BP-lowering trials. Moreover, intensive BP control can worsen limb perfusion and none of the trials adjudicated limb events.

**Methods:** Using data from a longitudinal registry of US Veterans, we identified 91,296 patients (>2.7 million BP values) with a new diagnosis of PAD during 2015-2020. Multivariable Cox regression models evaluated the association between longitudinal BP values (time-varying exposure) and risk of cardiovascular (CV: myocardial infarction, stroke) and limb (chronic limb threatening ischemia, major amputation) events.

**Results:** Mean age was 70.5 years, 97.3% were men and 18.5% were Black race. At baseline, the mean (SD) systolic BP was 133 (19) mm Hg and diastolic BP was 73 (11) mm Hg. Over a median 4.1 years, the incidence of CV and limb events was 4.6 per-100 patient-years and 5.0 per 100-patient-years, respectively. In adjusted Cox models, systolic BP ≥140 mm Hg was strongly associated with higher risk of CV events with increasing risk across BP categories (130-139 mm Hg [reference]; 140-149 mm Hg: hazard ratio [HR]: 1.28 [95%CI: 1.16-1.41]; 150-159 mm Hg: HR 1.54 [1.39-1.72]; ≥160 mm Hg: 2.22 [2.02-2.44]). However, there was no association between systolic BP below 130 mm Hg and risk of CV events. In contrast, the association between systolic BP and limb events was U-shaped with the highest hazard of limb events at both low (<110 mm Hg: HR: 1.61 [1.48-1.74]) and high (≥160 mm Hg: HR: 1.53 [1.41-1.67]) systolic BPs. This discordance between CV and limb events, especially at lower BP was more pronounced in those with diabetes and severe PAD. Similar findings were seen in the analysis of diastolic BP with CV and limb events.

**Conclusion:** Current recommendations for BP goal in PAD patients are associated with optimizing risk of CV events but associated with a higher risk of limb events. Our findings highlight the need to better define BP goals for PAD patients, especially those at an increased risk of limb events.

## Introduction

Based on multiple randomized controlled trials (RCTs) that have showed that intensive blood pressure (BP) control reduces cardiovascular (CV) events,^1–4^ current guidelines recommend a BP goal ≤130/80 mm Hg in all hypertensive patients (Class I), including those with peripheral artery disease (PAD).^5,6^ However, a benefit of intensive BP control in PAD is not established because, of more than 40,000 patients in the above RCTs, fewer than 2% (n=720) had PAD at baseline.^1–3^

In patients with PAD, the incidence of limb events such as chronic limb-threatening ischemia (CLTI) and major amputation is also high and often exceeds the incidence of CV events.^7,8^ Moreover, intensive BP control can worsen limb perfusion in the setting of lower extremity atherosclerosis.^9–13^ Yet, none of the landmark BP-lowering trials adjudicated CLTI or amputation, and therefore data regarding the effect of BP control on risk of limb events are limited. Finally, whether the association between BP and clinical events differs in subgroups at varying risk of CV and limb events (diabetes and severe PAD) remains unknown.

Accordingly, we leveraged the Peripheral Artery Disease Long-term Survival (PEARLS) registry, a national cohort of U.S. Veterans with incident PAD identified using a validated and highly accurate natural language processing (NLP) algorithm.^8,14^ The registry includes extensive clinical variables, repeated measures of BP, and long-term follow-up through linkage with multiple clinical and administrative databases, providing an opportunity to evaluate the association between longitudinal BP control and risk of both CV and limb events.

## Methods

This work was approved by the Institutional Review Board at UT Southwestern Medical Center and Research & Development Committees at Dallas, Iowa City and Tennessee Valley Health System Veterans Affairs (VA) Hospitals.

### Study Design

The design of the PEARLS registry has been previously described.^8^ Briefly, patients within the VA healthcare system with newly diagnosed PAD are identified based on abnormal ankle brachial index (ABI < 0.9) or a toe-brachial index (TBI <0.7) in either leg, with data extracted using a novel NLP system developed and validated by our research team.^14^ The positive predictive value (PPV) of the NLP system for PAD diagnosis (92.3%), is far superior to the reported PPV of International Classification of Disease codes.^15,16^ The PEARLS registry includes 103,748 Veterans with incident PAD between 2015-2020 with the index date defined as the date of first abnormal ABI or TBI.^8^ Clinical endpoints were identified using 1) VA claims from Corporate Data Warehouse (CDW), 2) services paid by VA for care at non-VA facilities in the fee-basis and consolidated datasets (CDS), and 3) 100% Medicare Part A (Inpatient), Part B (Outpatient) and Part C (Advantage) for those with Medicare insurance.

### Blood Pressure Data

Within the CDW, BP values are available in the Vital Sign domain. Only outpatient BP values were considered in the current analysis. Clinical staff in the VA (medical assistants, nurses) are instructed to measure BP after a 3-to-5-minute rest. We excluded BPs if they were a) missing systolic or diastolic values, b) systolic lower than diastolic c) systolic BP <60 mm Hg or >300 mm Hg and diastolic BP <30 mm Hg or >180 mm Hg. For multiple BP measurements on the same day, we included the lowest value.

### Study Cohort

We restricted our cohort to patients with at least 2 clinic visits during the 2 years prior to the index date, at least one BP reading during the year before the index date and within 1 year after the index date and those without any clinical event before their first BP reading after index date. We followed patients from index date to the time of first CV or limb event, death, or end of 5-years. In addition, patients with >3 years between BP readings were censored at the time of the last reading before this gap.

### Study Endpoints and Variables

Study endpoints included CV events (hospitalization for acute myocardial infarction or ischemic stroke) and limb events – CLTI (defined as ischemic rest pain, non-healing ulcers, or gangrene) or major limb amputation (ankle or above). All endpoints were identified within CDW and through linkage of data from the fee-basis, CDS and Medicare data as previously described.^8^ Because overall mortality was high and reliable cause-of-death information was unavailable, death was included as a censoring event.

The primary exposure was systolic and diastolic BP recorded during VA clinic visits. Since the association of BP with outcomes may be non-linear, we categorized systolic BP (mm Hg) as <110, 110-119, 120-129, 130-139 (reference), 140-149, 150-159, and ≥160 and diastolic BP (mm Hg) as <60, 60-69, 70-79 (reference), 80-89, ≥90. Study variables included age, sex, race, ethnicity, smoking, and co-morbidities - hypertension, diabetes, chronic kidney disease, chronic obstructive pulmonary disease, coronary artery disease, prior history of myocardial infarction, heart failure, arrhythmias, cancer, dementia, depression, chronic liver disease, anemia, valvular heart disease, weight loss, defined using Elixhauser and Charlson algorithm and included.^17^ We defined PAD severity as mild: 0.8 ≤ ABI ≤ 0.9 and 0.6 ≤ TBI ≤ 0.7; moderate: 0.5 ≤ ABI < 0.8 or 0.4 ≤ TBI < 0.6; severe: 0 ≤ ABI < 0.5 or 0 ≤ TBI < 0.4 in the worst limb.^18^

### Statistical Analysis

We compared patient characteristics across baseline systolic and diastolic BP categories using chi-square test for categorical variables, and linear regression for continuous variables.

To evaluate association of systolic and diastolic BP with CV and limb events, we constructed separate multivariable Cox proportional hazards regression models.^19^ The model was fitted in monthly increments and the proportion of time spent in each BP category was included as a time-varying exposure. Thus, as follow up accrued for each participation, the proportion of time spent in each category was updated based on new BP values. During months with no new BP values, data from the previous month were carried forward. Thus, an individual patient’s cumulative exposure to different BPs during follow up was captured as a time-varying exposure that was updated monthly from PAD diagnosis (index date) until the first event or end of follow up. Models were adjusted for age, sex, race, ethnicity, PAD severity, smoking, and all co-morbidities listed in the Study Endpoints and Variables Sections. In these models, death was considered a censoring event. Moreover, in analysis of CV risk, patients were not censored if they experienced a limb event before CV event and vice versa. A sandwich estimator was used to account for correlation between repeated BP values. The coefficients for each category were exponentiated to obtain a hazard ratio (HR) with 95% confidence intervals (CI). The proportionality hazards assumption was evaluated using an interaction of time with BP category, which was not significant.

To further quantify the marginal effect of exposure to each BP category, we also estimated the predicted incidence of CV and limb events over a 3-year period using recycled predictions, ^20^ with separate models for each event type. This measure represents the predicted incidence of CV (or limb event) for each BP category assuming that all patients in the cohort were exposed to the same BP category during follow-up while holding all other variables constant. Finally, we examined whether the association of BP with clinical events differed in subgroups with diabetes and severe PAD.

We conducted a range of sensitivity analyses to evaluate the robustness of our findings. First, in addition to death, we also censored patients if they experienced a limb event before a CV event (in analysis of CV risk), and vice versa. Second, given the high mortality and lack of reliable data on cause of death, we also considered death as a competing risk using a Fine-Gray model. However, inclusion of a time-varying exposure in a sub-distribution hazard model is not recommended because it interferes with the overall estimation of the cumulative incidence function and assessing the effect of the exposure on the incidence function leading to invalid inferences.^21^ Therefore, for this analysis we used a landmark approach, in which we fitted a set of conventional competing risk analysis (Fine-Gray) at 4 different intervals – index date, 1-year, 2-year and 3-years. At each landmark time, only patients who were alive and still at risk of experiencing the event were included, with time in each BP category until the landmark time included as a fixed covariate.

All analyses were performed using SAS version 9.4 (SAS Institute, Cary, North Carolina).

## Results

A total of 91,296 patients with >2.7 million BP values (mean: 30/patient) were included (Figure S1). At baseline, the mean (SD) systolic and diastolic BPs were 133 (19) mm Hg and 73 (11) mm Hg, respectively. During the first 12 months after index date, each patient spent a mean 3.0 to 4.9 months in the same systolic BP category and 3.5 to 6.1 months in the same diastolic BP category (Table S1), highlighting month-to-month variability in systolic and diastolic BP over time.

Table 1 shows patient characteristics in the overall cohort and stratified by baseline systolic BP. The mean (SD) age was 70.5 (9.0) years, 97.3% were men, 18.5% self-identified as Black race, and 3.0% as Hispanic ethnicity. More than 1 in 4 (27.3%) reported current smoking and 20.9% had severe PAD. The prevalence of co-morbidities especially diabetes (55.0%) and chronic kidney disease (23.3%) was high. Although many differences in demographics and co-morbidities across systolic and diastolic (Table S2) BP categories were statistically significant, most were small in magnitude.

**Table 1.** Characteristics of Study Cohort Based on Categories of Baseline Systolic Blood Pressure.

| *Variable* | *Overall*<br>*(N=91,296)* | *<110*<br>*(N=9,233)* | *110-119*<br>*(N=11,897)* | *120-129*<br>*(N=18,135)* | *130-139*<br>*(N=23,507)* | *140-149*<br>*(N=12,824)* | *150-159*<br>*(N=7,926)* | *≥160*<br>*(N=7,774)* | *P-*<br>*value* | *P-value*<br>*for trend* |
| --- | --- | --- | --- | --- | --- | --- | --- | --- | --- | --- |
| **Mean Age, yrs (SD)** | 70.5 (9.0) | 70.6 (9.1) | 70.3 (9.3) | 70.2 (9.0) | 70.4 (8.7) | 70.6 (8.9) | 70.9 (9.0) | 71.0 (9.0) | <0.001 | <0.001 |
| **Age Group** |  |  |  |  |  |  |  |  | <0.001 | <0.001 |
| 40-50 | 829 (0.9%) | 101 (1.1%) | 148 (1.2%) | 197 (1.1%) | 182 (0.8%) | 108 (0.8%) | 54 (0.7%) | 39 (0.5%) |  |  |
| 50-60 | 7,844 (8.6%) | 818 (8.9%) | 1,148 (9.6%) | 1,665 (9.2%) | 1,971 (8.4%) | 1,008 (7.9%) | 637 (8.0%) | 597 (7.7%) |  |  |
| 60-70 | 35,647 (39.0%) | 3,425 (37.1%) | 4,625 (38.9%) | 7,170 (39.5%) | 9,289 (39.5%) | 5,046 (39.3%) | 3,022 (38.1%) | 3,070 (39.5%) |  |  |
| 70-80 | 32,315 (35.4%) | 3,365 (36.4%) | 4,044 (34.0%) | 6,341 (35.0%) | 8,468 (36.0%) | 4,572 (35.7%) | 2,824 (35.6%) | 2,701 (34.7%) |  |  |
| ≥80 | 14,661 (16.1%) | 1,524 (16.5%) | 1,932 (16.2%) | 2,762 (15.2%) | 3,597 (15.3%) | 2,090 (16.3%) | 1,389 (17.5%) | 1,367 (17.6%) |  |  |
| **Sex** |  |  |  |  |  |  |  |  | 0.11 | 0.003 |
| Female | 2,479 (2.7%) | 269 (2.9%) | 359 (3.0%) | 499 (2.8%) | 623 (2.7%) | 345 (2.7%) | 196 (2.5%) | 188 (2.4%) |  |  |
| Male | 88,817 (97.3%) | 8,964 (97.1%) | 11,538 (97.0%) | 17,636 (97.2%) | 22,884 (97.3%) | 12,479 (97.3%) | 7,730 (97.5%) | 7,586 (97.6%) |  |  |
| **Race** |  |  |  |  |  |  |  |  | <0.001 | <0.001 |
| White | 68,232 (74.7%) | 7,121 (77.1%) | 9,048 (76.1%) | 13,632 (75.2%) | 17,642 (75.0%) | 9,470 (73.8%) | 5,766 (72.7%) | 5,553 (71.4%) |  |  |
| Black | 16,852 (18.5%) | 1,477 (16.0%) | 2,034 (17.1%) | 3,250 (17.9%) | 4,325 (18.4%) | 2,465 (19.2%) | 1,607 (20.3%) | 1,694 (21.8%) |  |  |
| Others | 2,068 (2.3%) | 209 (2.3%) | 265 (2.2%) | 441 (2.4%) | 508 (2.2%) | 281 (2.2%) | 181 (2.3%) | 183 (2.4%) |  |  |
| Unknown | 4,144 (4.5%) | 426 (4.6%) | 550 (4.6%) | 812 (4.5%) | 1,032 (4.4%) | 608 (4.7%) | 372 (4.7%) | 344 (4.4%) |  |  |
| **Ethnicity** |  |  |  |  |  |  |  |  | 0.51 | 0.79 |
| Hispanic | 2,764 (3.0%) | 264 (2.9%) | 347 (2.9%) | 546 (3.0%) | 716 (3.0%) | 378 (2.9%) | 265 (3.3%) | 248 (3.2%) |  |  |
| Non-Hispanic | 85,364 (93.5%) | 8,661 (93.8%) | 11,154 (93.8%) | 16,943 (93.4%) | 22,005 (93.6%) | 11,983 (93.4%) | 7,361 (92.9%) | 7,257 (93.3%) |  |  |
| Unknown | 3,168 (3.5%) | 308 (3.3%) | 396 (3.3%) | 646 (3.6%) | 786 (3.3%) | 463 (3.6%) | 300 (3.8%) | 269 (3.5%) |  |  |
| **PAD Severity** |  |  |  |  |  |  |  |  | <0.001 | <0.001 |
| Mild | 27,653 (30.3%) | 2,855 (30.9%) | 3,774 (31.7%) | 5,720 (31.5%) | 7,223 (30.7%) | 3,789 (29.5%) | 2,246 (28.3%) | 2,046 (26.3%) |  |  |
| Moderate | 44,584 (48.8%) | 4,399 (47.6%) | 5,778 (48.6%) | 8,836 (48.7%) | 11,635 (49.5%) | 6,277 (48.9%) | 3,898 (49.2%) | 3,761 (48.4%) |  |  |
| Severe | 19,059 (20.9%) | 1,979 (21.4%) | 2,345 (19.7%) | 3,579 (19.7%) | 4,649 (19.8%) | 2,758 (21.5%) | 1,782 (22.5%) | 1,967 (25.3%) |  |  |
| **Smoking Status** |  |  |  |  |  |  |  |  | <0.001 | <0.001 |
| Current | 24,905 (27.3%) | 2,524 (27.3%) | 3,220 (27.1%) | 5,013 (27.6%) | 6,443 (27.4%) | 3,467 (27.0%) | 2,126 (26.8%) | 2,112 (27.2%) |  |  |
| Former | 27,662 (30.3%) | 3,017 (32.7%) | 3,671 (30.9%) | 5,511 (30.4%) | 7,100 (30.2%) | 3,828 (29.9%) | 2,304 (29.1%) | 2,231 (28.7%) |  |  |
| Never | 11,601 (12.7%) | 1,065 (11.5%) | 1,558 (13.1%) | 2,386 (13.2%) | 2,935 (12.5%) | 1,650 (12.9%) | 1,013 (12.8%) | 994 (12.8%) |  |  |
| Unknown | 27,128 (29.7%) | 2,627 (28.5%) | 3,448 (29.0%) | 5,225 (28.8%) | 7,029 (29.9%) | 3,879 (30.2%) | 2,483 (31.3%) | 2,437 (31.3%) |  |  |
| **Co-morbidities** |  |  |  |  |  |  |  |  |  |  |
| Hypertension | 79,588 (87.2%) | 7,761 (84.1%) | 9,957 (83.7%) | 15,369 (84.7%) | 20,480 (87.1%) | 11,500 (89.7%) | 7,237 (91.3%) | 7,284 (93.7%) | <0.001 | <0.001 |
| Diabetes | 50,201 (55.0%) | 5,321 (57.6%) | 6,615 (55.6%) | 9,806 (54.1%) | 12,684 (54.0%) | 7,007 (54.6%) | 4,435 (56.0%) | 4,333 (55.7%) | <0.001 | 0.09 |
| CKD | 21,267 (23.3%) | 2,712 (29.4%) | 2,805 (23.6%) | 3,897 (21.5%) | 4,906 (20.9%) | 2,993 (23.3%) | 1,881 (23.7%) | 2,073 (26.7%) | <0.001 | 0.01 |
| COPD | 32,707 (35.8%) | 4,180 (45.3%) | 4,682 (39.4%) | 6,546 (36.1%) | 8,007 (34.1%) | 4,271 (33.3%) | 2,601 (32.8%) | 2,420 (31.1%) | <0.001 | <0.001 |
| Arrhythmia | 27,323 (29.9%) | 4,120 (44.6%) | 4,102 (34.5%) | 5,427 (29.9%) | 6,222 (26.5%) | 3,425 (26.7%) | 2,084 (26.3%) | 1,943 (25.0%) | <0.001 | <0.001 |
| Heart failure | 20,378 (22.3%) | 3,556 (38.5%) | 3,177 (26.7%) | 3,961 (21.8%) | 4,241 (18.0%) | 2,408 (18.8%) | 1,509 (19.0%) | 1,526 (19.6%) | <0.001 | <0.001 |
| CAD | 39,846 (43.6%) | 5,189 (56.3%) | 5,744 (48.2%) | 7,928 (43.7%) | 9,580 (40.8%) | 5,112 (39.9%) | 3,204 (40.3%) | 3,089 (39.8%) | <0.001 | <0.001 |
| CVD | 21,741 (23.8%) | 2,491 (27.0%) | 2,809 (23.6%) | 4,133 (22.8%) | 5,373 (22.9%) | 2,986 (23.3%) | 1,945 (24.5%) | 2,004 (25.8%) | <0.001 | 0.53 |
| Cancer | 14,623 (16.0%) | 1,761 (19.1%) | 1,973 (16.6%) | 2,806 (15.5%) | 3,600 (15.3%) | 2,079 (16.2%) | 1,252 (15.8%) | 1,152 (14.8%) | <0.001 | <0.001 |
| Dementia | 4,146 (4.5%) | 603 (6.5%) | 649 (5.5%) | 794 (4.4%) | 873 (3.7%) | 538 (4.2%) | 331 (4.2%) | 358 (4.6%) | <0.001 | <0.001 |
| Depression | 28,438 (31.1%) | 3,359 (36.4%) | 4,086 (34.3%) | 5,832 (32.2%) | 6,841 (29.1%) | 3,771 (29.4%) | 2,324 (29.3%) | 2,225 (28.6%) | <0.001 | <0.001 |
| Chronic liver disease | 8,608 (9.4%) | 1,077 (11.7%) | 1,180 (9.9%) | 1,635 (9.0%) | 2,100 (8.9%) | 1,157 (9.0%) | 731 (9.2%) | 728 (9.4%) | <0.001 | <0.001 |
| Obesity | 22,771 (24.9%) | 2,466 (26.7%) | 3,078 (25.9%) | 4,561 (25.2%) | 5,915 (25.2%) | 3,149 (24.6%) | 1,891 (23.9%) | 1,711 (22.0%) | <0.001 | <0.001 |
| Anemia | 9,838 (10.8%) | 1,453 (15.7%) | 1,390 (11.7%) | 1,898 (10.5%) | 2,171 (9.2%) | 1,300 (10.1%) | 801 (10.1%) | 825 (10.6%) | <0.001 | <0.001 |
| Valve disease | 9,549 (10.5%) | 1,413 (15.3%) | 1,454 (12.2%) | 1,892 (10.4%) | 2,111 (9.0%) | 1,203 (9.4%) | 752 (9.5%) | 724 (9.3%) | <0.001 | <0.001 |
| Weight loss | 6,480 (7.1%) | 1,058 (11.5%) | 999 (8.4%) | 1,244 (6.9%) | 1,338 (5.7%) | 847 (6.6%) | 484 (6.1%) | 510 (6.6%) | <0.001 | <0.001 |

**Abbreviations:** CAD: coronary artery disease; CVD: cerebrovascular disease; CKD: chronic kidney disease; COPD: chronic obstructive pulmonary disease; PAD: peripheral artery disease; SD: standard deviation

Over a median 4.1 years, 15,500 patients experienced a CV event (incidence: 4.6 per-100 patient-years), 16,314 experienced a limb event (5.0 per-100 patient-years) and 21,736 patients died (7.2 per-100 patient-years). The incidence of limb events was higher than CV events in patients with diabetes (6.2 vs. 5.4 per-100 patient-years) and severe PAD (10.5 vs. 5.9 per-100 patient-years) but similar in older patients (5.1 vs. 5.0 per-100 patient years, Table 2).

**Table 2.** Unadjusted Incidence of Cardiovascular and Limb Event Overall and Within Subgroups. The overall number and incidence rate, per-100 patient years (parenthesis) are reported for the overall cohort and stratified by age (<70 vs. ≥70 years), presence of diabetes, and severity of PAD

|  | *Incidence Rate (per-100 patient years)* |  |  |
| --- | --- | --- | --- |
|  | *Cardiovascular Event* | *Limb Event* | *Death* |
| **Overall** | **15,500 (4.6)** | **16,314 (5.0)** | **21,736 (7.2)** |
| Diabetes |  |  |  |
| No | 5,861 (3.7) | 5,666 (3.7) | 9,226 (6.4) |
| Yes | 9,639 (5.4) | 10,648 (6.2) | 12,510 (8.0) |
| Severe PAD |  |  |  |
| No | 11,748 (4.3) | 10,440 (3.9) | 17,051 (6.8) |
| Yes | 3,752 (5.9) | 5,874 (10.5) | 4,685 (9.1) |

**Table 3.** Risk-adjusted Hazard Ratio of Cardiovascular and Limb Events Across Categories of Systolic and Diastolic Blood Pressure. The hazard ratios (95% confidence intervals) are reported for each category of systolic blood pressure (reference: 130-139 mm Hg) and diastolic blood pressure (reference: 70-79 mm Hg) from a multivariable Cox regression model with time-varying BP and adjusting for age, sex, race, ethnicity, severity of PAD, smoking and all-comorbidities. Death was included as a censoring event.

|  | *Systolic Blood Pressure (mm Hg)* |  |  |  |  |  |  |
| --- | --- | --- | --- | --- | --- | --- | --- |
|  | **<110** | **110-120** | **120-129** | **130-139** | **140-149** | **150-159** | **≥160** |
| Cardiovascular Event | 1.11<br>(1.00-1.23) | 0.92<br>(0.83-1.02) | 0.94<br>(0.86-1.03) | Reference | 1.28<br>(1.16-1.41) | 1.54<br>(1.39-1.72) | 2.22<br>(2.02-2.44) |
| Limb Event | 1.61<br>(1.48-1.74) | 1.26<br>(1.16-1.36) | 1.16<br>(1.08-1.25) | Reference | 1.23<br>(1.13-1.33) | 1.30<br>(1.19-1.43) | 1.53<br>(1.41-1.67) |
|  | *Diastolic Blood Pressure (mm Hg)* |  |  |  |  |  |  |
|  | **<60** | **60-69** | **70-79** | **80-89** | **≥90** |  |  |
| Cardiovascular Event | 1.16<br>(1.06-1.26) | 1.07<br>(0.99-1.15) | Reference | 1.11<br>(1.02-1.20) | 1.92<br>(1.72-2.15) |  |  |
| Limb Event | 1.30<br>(1.21-1.40) | 1.11<br>(1.05-1.17) | Reference | 0.91<br>(0.85-0.97) | 1.21<br>(1.10-1.33) |  |  |

In multivariable Cox models with time-varying BP, there was a non-linear association between systolic BP and risk of CV events. Compared to the 130-139 mm Hg (reference), there was no increase in hazard of CV events in systolic BP categories below 130 mm Hg (Figure 1 and Table 2). However, above 140 mm Hg, there was a significant positive association between systolic BP and the hazard of CV events, with more than 2-fold increased hazard for systolic BP ≥ 160 mm Hg (hazard ratio [HR]: 2.22 [95% CI: 2.02-2.44]). In contrast, the association between systolic BP and limb events was U-shaped. Compared to the 130-139 mm Hg category, the hazard of limb events was significantly higher in categories below and above that threshold, with the highest risk in patients with very low (<110 mm Hg: Hg: HR 1.61; 95% CI: 1.48-1.74) and very high (≥160 mm Hg: HR 1.53; 95% CI: 1.41-1.67; Figure 1A and Table 2) systolic BP. In absolute terms, the 3-year predicted incidence of CV events was highest (22.5%) if systolic BP was ≥160 mm Hg for all patients holding other variables constant (Figure 2A and Table S3). In contrast, the 3-year predicted incidence of limb events was highest for systolic BP <110 mm Hg (18.1%) and ≥160 mm Hg (17.4%), whereas the lowest incidence (11.8%) was in the 130-139 mm Hg category (Figure 2A and Table S3).

**Figure 1.**
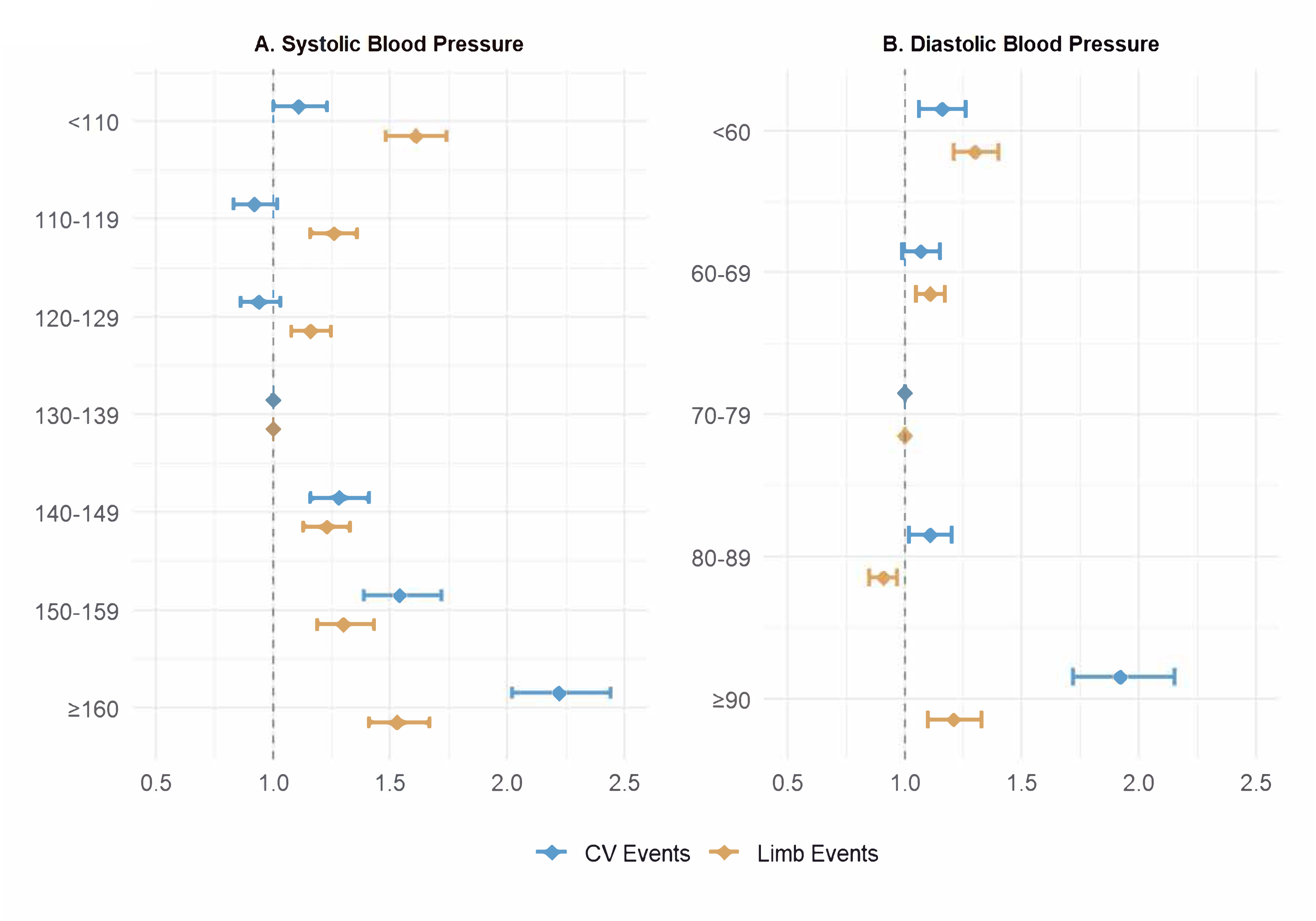
Association of Systolic and Diastolic BP with Risk of Cardiovascular and Limb Events. The forest plot shows risk-adjusted hazard ratio for each category of systolic BP (Panel A) and diastolic BP (Panel B) using a multivariable Cox regression model with time-varying BP and adjusting for age, sex, race, ethnicity, severity of PAD, smoking and all-comorbidities. The reference category is 130-139 mm Hg for systolic BP and 70-79 mm Hg for diastolic BP.

**Figure 2.**
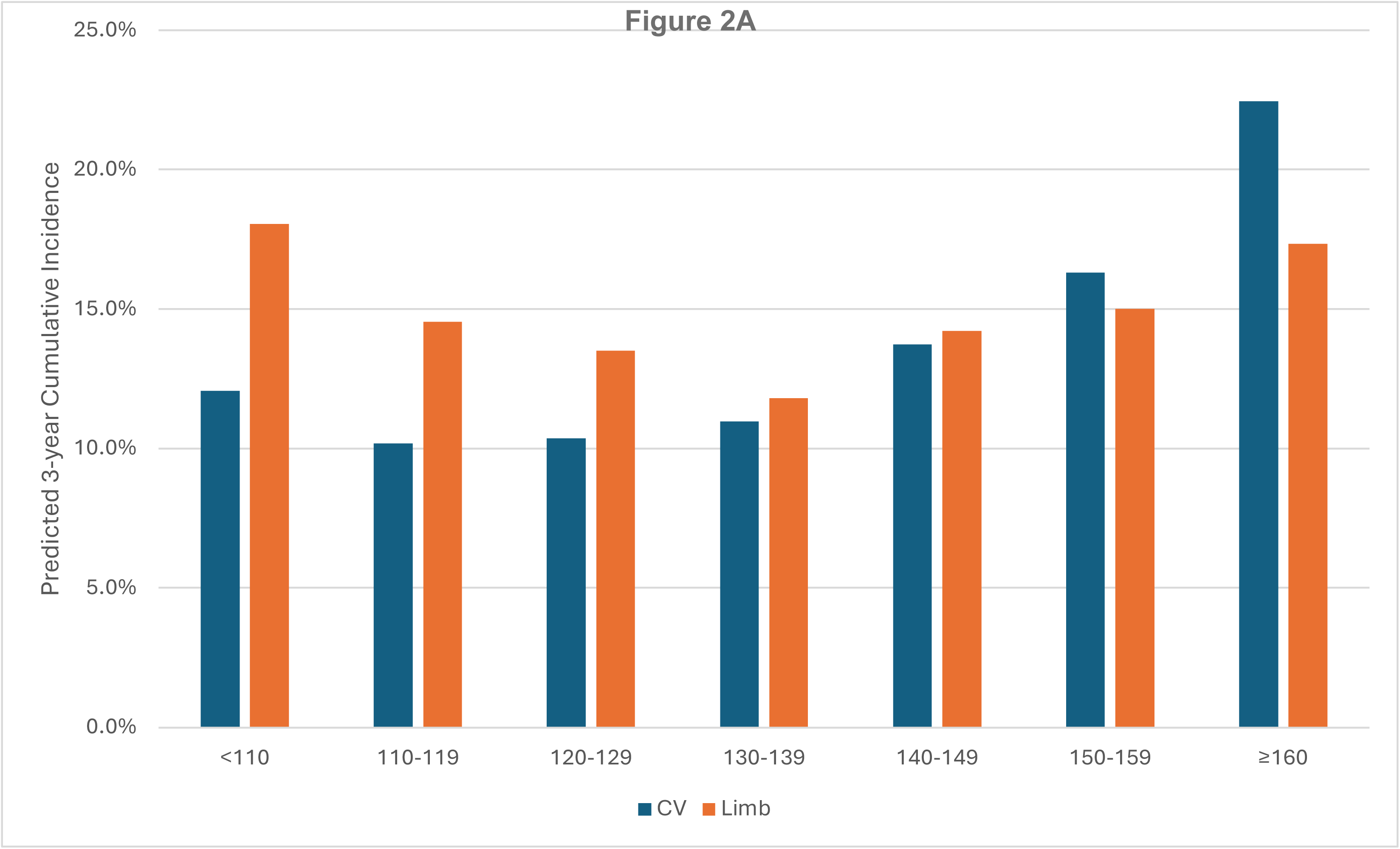
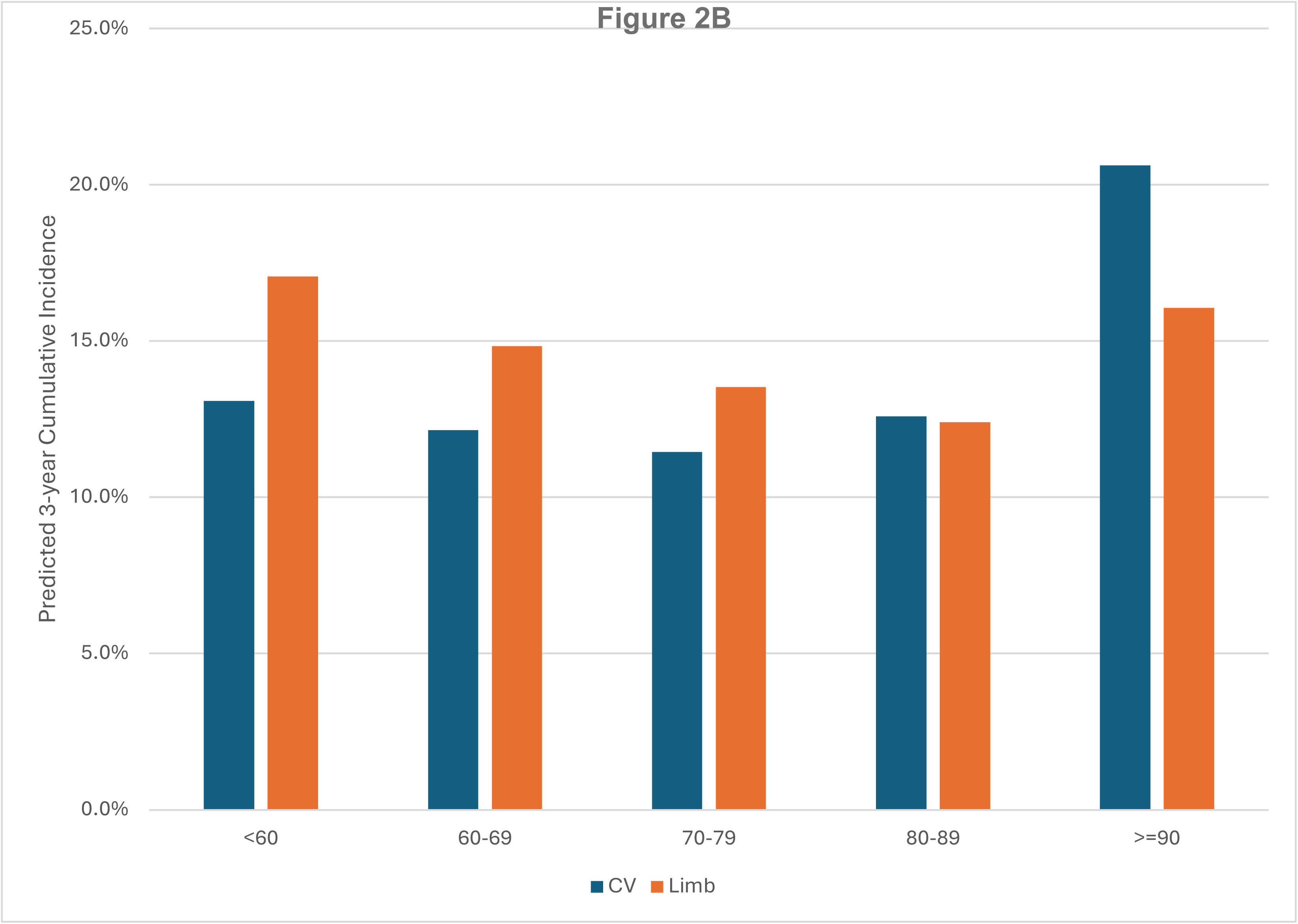
Predicted 3-year Incidence of Cardiovascular & Limb Events Across Categories of Systolic and Diasatolic Blood Pressure. We applied recycled predictions to the estimates from the Cox to quantify the marginal effect of systolic BP (Panel A) and diastolic BP (Panel B) on the 3-year incidence of cardiovascular and limb events. This method assumes identical BP for all patients in the cohort while holding all other variables constant. For example, if the entire cohort maintained a systolic BP of 130-139 mm Hg, the 3-year predicted incidence of CV events would be 11.0% and limb events would be 11.8%.

The association between diastolic BP with risk of clinical events was similar. Compared to reference (70-79 mm Hg), there was a small increase in risk of CV events with both lower and higher diastolic BPs, except diastolic BP ≥90 mm Hg which was associated with a 92% higher hazard of CV events (HR: 1.92 [95% CI: 1.72-2.15], Figure 2B and Table 2). A modest U-shaped association was also present between diastolic BP and limb events, with a 30% and 21% increase in hazard of limb events in those with diastolic BP <60 mm hg and ≥90 mm Hg, respectively. The lowest hazard of limb events was noted in the diastolic BP 80-89 mm Hg. In absolute terms, 3-year predicted incidence of CV and limb events was lowest in the 70-79 mm Hg (11.5%) and the 80-89 mm Hg (12.4%) diastolic BP category, respectively (Figure 2B and Table S3).

The discordance between systolic BP and its association CV vs. limb events, especially at lower BPs was more pronounced in those with diabetes (Table S4 and Figure S2 Panel A) and severe PAD (Table S5 and Figure S2 Panel B), groups with a higher incidence of limb events compared to CV events. For example, in severe PAD patients, the predicted cumulative 3-year incidence of limb events was nearly double the incidence of CV events in all systolic BP categories below 140 mm Hg (Table S5).

In sensitivity analysis, we found that the association of BP with CV and limb events remained largely unchanged when we censored patients with limb events prior to CV events (in analysis of CV events) and CV events prior to limb events (in analysis of limb events, Table S7). Our findings were also consistent in analysis that accounted for death as a competing risk (Table S8).

## Discussion

In this study of >90,000 Veterans with incident PAD, we found that the association of BP with clinical outcomes differed by type of event (CV vs. limb). Consistent with RCTs, systolic BPs above 130 mm Hg and diastolic BPs above 80 mm Hg were associated with higher risk of CV events. In contrast, the association between BP and limb events, was U-shaped, with elevated risk observed both above and below these thresholds. In fact, the lowest risk of limb events was observed in patients with systolic BP of 130-139 mm Hg and a diastolic BP of 80-89 mm Hg—which is above the threshold of 130/80 mm Hg recommended in PAD guidelines.^5^ Moreover, limb events exceeded CV events in our cohort, especially in those with diabetes and severe PAD. Given their systematic under-representation in landmark BP-lowering trials, a BP goal recommendation of ≤130/80 mm Hg for patients with PAD may need to be reevaluated, especially in those at highest risk of limb events.

Multiple RCTs over the past decade have shown a significant reduction in risk of CV events with intensive BP control in patients with hypertension, including older patients and those with diabetes.^1–4^ Based on these studies, a BP goal of <130/80 is recommended (Class I) for all hypertensive patients, including PAD.^5,6^ However, only 72 of 11,255 (0.6%) patients in the Effects of Intensive Systolic Blood Pressure Lowering Treatment in Reducing Risk of Vascular Events (ESPRIT),^1^ 145 of 12,821 (1.1%) in Blood Pressure Control in Diabetes (BP-ROAD),^2^ and 503 of 9361 (5.3%) in Systolic Blood Pressure Intervention Trial (SPRINT)^3^ had PAD at baseline. Subgroup analysis stratified by PAD status have only been reported in SPRINT which found no heterogeneity in the effect of intensive BP control on CV risk.^22^ Consistent with this finding, we also found a linear association between systolic BP above 140 mm Hg and CV events in >90,000 patients with PAD.

However, our finding of the U-shaped association between BP and limb events fills an important knowledge gap because none of the prior RCTs adjudicated limb events. Consistent with prior studies,^7,8^ major amputation and CLTI were more common than CV events in our PAD cohort and limb events can have devastating consequences. The resulting loss of mobility and functional decline can sharply erode quality of life and contribute to the excess mortality in PAD patients.^23,24^ In SPRINT, limb events such as CLTI or major amputation were not adjudicated. However, a post hoc analysis found that the incidence of peripheral artery surgery in the intensive BP control arm was lower compared to the standard treatment arm.^25^ In contrast, our PAD cohort experienced 16,325 CLTI or major amputation events with an overall incidence (5.0 per-100 patient-years) which exceeded the incidence of CV events, especially in those with diabetes (6.4 per-100 patient-years) and severe PAD (10.9 per-100 patient years). Thus, while a strategy of intensive BP control may optimize the risk of CV events, a systolic BP <130 mm Hg was associated with a stepwise increase in the risk of limb events highlighting important trade-offs. These trade-offs were even more important in those with diabetes and severe PAD, two groups at the highest risk of limb events. In fact, the lowest risk of limb events in our study was observed in the 130-139 mm Hg category for systolic and 80-89 mm Hg category for diastolic BP, both of which are above ≤130/80 mm Hg goal recommended in PAD guidelines.^5^

Our findings are consistent with the possibility that impaired limb perfusion at lower BPs increases the risk of limb-related complications.^12^ Compared with cerebral and coronary circulation, autoregulatory mechanisms are less potent in maintaining perfusion in the skeletal muscle and limbs.^13^ A J-shaped association between systolic BP and limb events was also reported in post-hoc analysis of the Antihypertensive and Lipid-Lowering Treatment to Prevent Heart Attack Trial (ALLHAT). ^26^ However, in contrast to our study, ALLHAT enrolled very few patients with PAD at baseline. Thus, nearly all of the limb events in that study represented new-onset PAD. Moreover, the definition of limb events in ALLHAT (PAD-related hospitalization, revascularization, medications and death) lacked specificity, and did not include amputation.^27^ In contrast, Fudim et al found increased risk of limb events with systolic BP >125 mm Hg, but no excess risk at lower BP.^28^ The incidence of major amputation and CLTI in this study was exceedingly low and >80% of limb events were revascularization procedures which are discretionary and may not always reflect progression of PAD. Finally, in a recent RCT of hypertensive patients with end-stage renal disease, intensive BP control (110-140 mmHg) resulted in a 3-fold increase in risk of vascular access thrombosis highlighting the effect of intensive BP control on vascular events in a group that has a high prevalence of PAD.^27^ Collectively, these data highlight a need to better understand whether intensive BP control is beneficial or harmful in PAD patients especially with regards to risk of limb events.

There are several strengths of our study that deserve emphasis. First, identification of PAD in our study was based on a validated NLP system that used ABI and TBI values which was shown to be far superior to diagnosis codes (positive predictive value of 92.3%).^8,14^ Second, longitudinal BP data from clinic visits was available, facilitating the use of a time-varying model. Third, linkage of our cohort with Fee-basis, CDS and 100% Medicare claims (Part A-D) is a key strength that allowed us to capture clinical events for Veterans who also visit non-VA provides for their health needs. We previously showed that the addition of these linked data sources increased the yield of CV events by more than 300%.^8^

Our findings should be interpreted in context of several limitations. First, our exposure was based on office BP which may be higher than BP in a research setting.^29^ However, this type of misclassification cannot account for the differential association of BP with CV and limb events and our findings reflect BP values routinely used to make treatment decisions. Second, despite the high mortality in our cohort, we lacked information on cause of death. To address this, we included death as a competing risk in sensitivity analyses and found that the overall association of BP with CV and limb events was similar. Third, although our analytic approach considered BP as a time-varying exposure and adjusted for a range of possible confounders, there is potential for residual confounding. Fourth, although most demographic characteristics were well represented, women and Hispanics comprised <3%, limiting the generalizability of our findings to these important subgroups.

In conclusion, we found a disparate relationship between BP and CV and limb events. While higher BP was associated with a stepwise increased risk of CV events, the association with limb events was non-linear with increased risk at both high and low BP. Our findings highlight important trade-offs in optimizing risk associated with intensive BP control in patients with PAD.

## Data Availability

Data supporting this study are available from the Department of Veterans Affairs to qualified researchers with appropriate permissions to access data on Veterans.

## REFERENCES

1. Liu J, Li Y, Ge J, et al. Lowering systolic blood pressure to less than 120 mm Hg versus less than 140 mm Hg in patients with high cardiovascular risk with and without diabetes or previous stroke: an open-label, blinded-outcome, randomised trial. Lancet 2024;404(10449):245–255. DOI: 10.1016/S0140-6736(24)01028-6.

2. Bi Y, Li M, Liu Y, et al. Intensive Blood-Pressure Control in Patients with Type 2 Diabetes. N Engl J Med 2024. DOI: 10.1056/NEJMoa2412006.

3. Group SR, Wright JT, Jr., Williamson JD, et al. A Randomized Trial of Intensive versus Standard Blood-Pressure Control. N Engl J Med 2015;373(22):2103–16. DOI: 10.1056/NEJMoa1511939.

4. Zhang W, Zhang S, Deng Y, et al. Trial of Intensive Blood-Pressure Control in Older Patients with Hypertension. N Engl J Med 2021;385(14):1268–1279. DOI: 10.1056/NEJMoa2111437.

5. Gornik HL, Aronow HD, Goodney PP, et al. 2024 ACC/AHA/AACVPR/APMA/ABC/SCAI/SVM/SVN/SVS/SIR/VESS Guideline for the Management of Lower Extremity Peripheral Artery Disease: A Report of the American College of Cardiology/American Heart Association Joint Committee on Clinical Practice Guidelines. Circulation 2024;149(24):e1313–e1410. DOI: 10.1161/CIR.0000000000001251.

6. Whelton PK, Carey RM, Aronow WS, et al. 2017 ACC/AHA/AAPA/ABC/ACPM/AGS/APhA/ASH/ASPC/NMA/PCNA Guideline for the Prevention, Detection, Evaluation, and Management of High Blood Pressure in Adults: Executive Summary: A Report of the American College of Cardiology/American Heart Association Task Force on Clinical Practice Guidelines. Hypertension 2018;71(6):1269–1324. DOI: 10.1161/HYP.0000000000000066.

7. Kumbhani DJ, Steg PG, Cannon CP, et al. Statin therapy and long-term adverse limb outcomes in patients with peripheral artery disease: insights from the REACH registry. Eur Heart J 2014;35(41):2864–72. DOI: 10.1093/eurheartj/ehu080.

8. Girotra S, Li Q, Vaughan-Sarrazin M, et al. Long-Term Outcomes of Peripheral Artery Disease in Veterans: Analysis of the Peripheral Artery Disease Long-Term Survival Study (PEARLS). J Am Heart Assoc 2025;14(7):e038403. DOI: 10.1161/JAHA.124.038403.

9. Hirsch AT, Criqui MH, Treat-Jacobson D, et al. Peripheral arterial disease detection, awareness, and treatment in primary care. JAMA 2001;286(11):1317–24. (http://www.ncbi.nlm.nih.gov/pubmed/11560536).

10. Willey J, Mentias A, Vaughan-Sarrazin M, McCoy K, Rosenthal G, Girotra S. Epidemiology of lower extremity peripheral artery disease in veterans. Journal of vascular surgery 2018;68(2):527–535 e5. DOI: 10.1016/j.jvs.2017.11.083.

11. Franklin H, Rajan M, Tseng CL, Pogach L, Sinha A, Mph M. Cost of lower-limb amputation in U.S. veterans with diabetes using health services data in fiscal years 2004 and 2010. J Rehabil Res Dev 2014;51(8):1325–30. DOI: 10.1682/JRRD.2013.11.0249.

12. Fudim M, Jones WS. New Curveball for Hypertension Guidelines? Circulation 2018;138(17):1815–1818. DOI: 10.1161/CIRCULATIONAHA.118.036409.

13. Johnson PC. Autoregulation of blood flow. Circ Res 1986;59(5):483–95. DOI: 10.1161/01.res.59.5.483.

14. Friberg JE, Qazi AH, Boyle B, et al. Ankle- and Toe-Brachial Index for Peripheral Artery Disease Identification: Unlocking Clinical Data Through Novel Methods. Circ Cardiovasc Interv 2022;15(3):e011092. DOI: 10.1161/CIRCINTERVENTIONS.121.011092.

15. Hong Y, Sebastianski M, Makowsky M, Tsuyuki R, McMurtry MS. Administrative data are not sensitive for the detection of peripheral artery disease in the community. Vasc Med 2016;21(4):331–6. DOI: 10.1177/1358863X16631041.

16. Fan J, Arruda-Olson AM, Leibson CL, et al. Billing code algorithms to identify cases of peripheral artery disease from administrative data. Journal of the American Medical Informatics Association : JAMIA 2013;20(e2):e349–54. DOI: 10.1136/amiajnl-2013-001827.

17. Quan H, Sundararajan V, Halfon P, et al. Coding algorithms for defining comorbidities in ICD-9-CM and ICD-10 administrative data. Med Care 2005;43(11):1130–9. (https://www.ncbi.nlm.nih.gov/pubmed/16224307).

18. Aboyans V, Criqui MH, Abraham P, et al. Measurement and interpretation of the ankle-brachial index: a scientific statement from the American Heart Association. Circulation 2012;126(24):2890–909. DOI: 10.1161/CIR.0b013e318276fbcb.

19. Fine JP, and Gray RJ. A Proportional Hazards Model for the Subdistribution of a Competing Risk. Journal of the American Statistical Association 1999;94(446):496–509. DOI: 10.1080/01621459.1999.10474144.

20. Basu A, Rathouz PJ. Estimating marginal and incremental effects on health outcomes using flexible link and variance function models. Biostatistics 2005;6(1):93–109. DOI: 10.1093/biostatistics/kxh020.

21. Austin PC, Latouche A, Fine JP. A review of the use of time-varying covariates in the Fine-Gray subdistribution hazard competing risk regression model. Stat Med 2020;39(2):103–113. DOI: 10.1002/sim.8399.

22. Frary JMC, Pareek M, Byrne C, et al. Intensive blood pressure control appears to be effective and safe in patients with peripheral artery disease: the Systolic Blood Pressure Intervention Trial. Eur Heart J Cardiovasc Pharmacother 2021;7(3):e38–e40. DOI: 10.1093/ehjcvp/pvaa102.

23. Farber A. Chronic Limb-Threatening Ischemia. N Engl J Med 2018;379(2):171-180. DOI: 10.1056/NEJMcp1709326.

24. Menard MT, Farber A, Powell RJ, et al. Quality of Life in Patients With Chronic Limb-Threatening Ischemia Treated With Revascularization. Circulation 2024;149(16):1241–1253. DOI: 10.1161/CIRCULATIONAHA.123.065277.

25. Krogager ML, Pareek M, Kragholm KH, et al. Intensive vs. standard blood pressure control and vascular procedures: insights from the Systolic Blood Pressure Intervention Trial (SPRINT). Eur Heart J Cardiovasc Pharmacother 2021;7(3):e35–e37. DOI: 10.1093/ehjcvp/pvaa093.

26. Itoga NK, Tawfik DS, Lee CK, Maruyama S, Leeper NJ, Chang TI. Association of Blood Pressure Measurements With Peripheral Artery Disease Events. Circulation 2018;138(17):1805–1814. DOI: 10.1161/CIRCULATIONAHA.118.033348.

27. Miskulin DC, Gassman J, Schrader R, et al. BP in Dialysis: Results of a Pilot Study. J Am Soc Nephrol 2018;29(1):307–316. DOI: 10.1681/ASN.2017020135.

28. Fudim M, Hopley CW, Huang Z, et al. Association of Hypertension and Arterial Blood Pressure on Limb and Cardiovascular Outcomes in Symptomatic Peripheral Artery Disease: The EUCLID Trial. Circulation Cardiovascular quality and outcomes 2020;13(9):e006512. DOI: 10.1161/CIRCOUTCOMES.120.006512.

29. Drawz PE, Agarwal A, Dwyer JP, et al. Concordance Between Blood Pressure in the Systolic Blood Pressure Intervention Trial and in Routine Clinical Practice. JAMA Intern Med 2020;180(12):1655–1663. DOI:10.1001/jamainternmed.2020.5028.

